# Leveraging Language Embeddings from EMA Surveys to Predict Perceived Social Isolation among Stroke Survivors

**DOI:** 10.1101/2025.07.17.25331714

**Authors:** Yunda Liu, Alex W. K. Wong, Mandy W. M. Fong, Christopher L. Metts, Yun Shi, Sunghoon Ivan Lee

**Affiliations:** Manning College of Information and Computer Sciences, University of Massachusetts Amherst, Amherst, MA 01003 USA; Center for Rehabilitation Outcomes Research, Shirley Ryan AbilityLab, IL 60611 USA; Department of Physical Medicine and Rehabilitation, Northwestern University Feinberg School of Medicine, IL 60611 USA; Department of Medical Social Sciences, Northwestern University Feinberg School of Medicine, IL 60611 USA; Michigan Avenue Neuropsychologists, IL 60601 USA; Department of Pathology and Laboratory Medicine, Medical University of South Carolina, SC 29425 USA; Department of Neurology, New York University Grossman School of Medicine, NY 10017 USA

**Keywords:** Stroke, social isolation, loneliness, ecological momentary assessment, smartphone, natural language processing

## Abstract

Perceived social isolation (PSI) significantly affects the emotional well-being of stroke survivors, necessitating effective monitoring and prediction for timely, targeted interventions. While Ecological Momentary Assessment (EMA) has been increasingly used to identify precursor characteristics of PSI, existing prediction methods rely on handcrafted features, which often fail to capture the semantic richness and contextual relationships among survey questions. In this study, we propose a novel approach to predict PSI by processing structured EMA data with language embeddings. A total of 11,802 EMA surveys were collected from 218 stroke survivors, the largest dataset of its kind in social isolation research for this population. Language embeddings were extracted from the structured EMA surveys using a pre-trained language model. These embeddings were then processed by training an autoencoder to generate compact latent representations, which were used for the downstream PSI prediction. Our findings show that the proposed approach achieves accurate PSI prediction, with a weighted F_1_ score of 0.84 and a weighted AUPRC of 0.92, outperforming traditional handcrafted features. Furthermore, by leveraging only three carefully selected questions, our method can optimize a trade-off between validity and usability. This study demonstrates an efficient method for real-time monitoring of psychosocial outcomes in stroke survivors, with potential implications for early intervention and personalized care.

## I. INTRODUCTION

**S**TROKE can lead to physical and cognitive impairments that significantly disrupt survivors’ daily functioning [1]. These disruptions often result in a profound disconnection from pre-stroke self-identity and environments, driven by challenges such as job loss and role changes, leading to reduced social and community engagement [2]. While rehabilitation seeks to improve motor and cognitive functioning, many stroke survivors face persistent psychosocial challenges [1], which often intensify during the later stages of recovery [3]. Consequently, social isolation is common, affecting nearly half of stroke survivors 12 months post-stroke [4], [5]. Beyond causing emotional distress [6], [7], social isolation is a risk factor for adverse clinical outcomes, including poorer functional recovery, greater functional decline, and an increased risk of recurrent stroke [8]. Thus, there is a critical need to monitor and predict perceived social isolation (PSI) for timely, targeted interventions.

To enable continuous monitoring, Ecological Momentary Assessment (EMA) has been widely used as an effective tool for collecting behavioral and psychosocial data in naturalistic environments [9]. EMA involves repeated real-time assessments, aiming to reduce recall bias and enhance ecological validity by capturing fluctuations in individuals’ experiences and behaviors in daily life [9], [10]. Moreover, with advancements in mobile technologies, EMA has become increasingly flexible and scalable, making it practical for longitudinal monitoring of psychosocial outcomes [11]. Prior research has demonstrated that smartphone-based EMA can accurately measure mood states, post-stroke symptoms, and daily behaviors among stroke survivors, offering valuable insights into their real-world experiences and recovery trajectories [12]–[14].

To predict PSI using EMA data, existing approaches typically employ machine learning models trained on manually crafted, discrete features from patients’ responses [15]–[17]. These features, often derived from simple statistical metrics (e.g., mean, standard deviation) [18] and categorical encoding [19], have shown promise in capturing basic response patterns. However, such approaches often overlook the nuanced semantic meanings embedded in natural language and fail to model complex inter-question relationships within survey data. To address these limitations, recent studies—though limited— have begun applying natural language processing (NLP) to EMA data analysis [20], [21]. Yet, these emerging approaches either sacrifice semantic precision or require free-text inputs, which can be burdensome for patients during frequent, daily assessments, ultimately limiting their near real-time predictive utility [22].

In this work, we present a data analytic pipeline for predicting PSI among stroke survivors by leveraging NLP techniques to extract contextualized language embeddings from structured EMA data. Building on the recent success in language models, we hypothesized that the use of language embeddings enables richer, more comprehensive representations of stroke survivors’ momentary experiences and mental states, thereby enhancing PSI prediction accuracy. To test this, we utilized a dataset of 11,082 EMA surveys collected from 218 stroke survivors, the largest dataset of its kind in social isolation research for this population. We extracted embeddings using a pre-trained language model and trained an autoencoder to generate compact latent representations, which were then used as input for a downstream PSI prediction model. Furthermore, we hypothesized that information across multiple EMA surveys per participant can capture temporal patterns and improve predictive performance. Our promising results highlight the potential for developing low-burden, scalable tools for continuous monitoring of psychosocial well-being in stroke survivors, paving the way for personalized and timely interventions in real-world settings.

## II. METHOD

### A. Study Participants

We utilized a dataset of 218 stroke survivors (95 female, 59.79 ± 11.74 years old; mean ± standard deviation) in this study [23]. Study participants were recruited from a single hospital database between October 2018 and January 2021. To be eligible, participants must 1) be at least 18 years old, 2) be fluent in English, 3) exhibit mild-to-moderate stroke severity with the National Institutes of Health Stroke Scale (NIHSS) score of 13 or lower, 4) have experienced an ischemic or hemorrhagic stroke at least 3 months prior to the study, and 5) have no or minor pre-stroke disabilities. The study was approved by the ethics committees of Washington University in Saint Louis (#201704024) and Northwestern University (#STU00215308).

### B. Data Collection

During the study, participants were instructed to complete 5 EMA surveys per day for 14 consecutive days, totaling up to 70 surveys per participant. However, due to non-compliance and other barriers, each participant completed an average of 54.14 *±* 15.79 surveys throughout the study period.

Participants completed EMA surveys using a mobile application installed on either 1) an iPod Touch (sixth generation, iOS 11) provided by the research team or 2) their own iPhone. During the study, the mobile application prompted participants to complete daily surveys at random intervals of approximately 2.5 hours within specific time slots: 8:00 AM to 10:00 AM, 11:00 AM to 1:00 PM, 2:00 PM to 4:00 PM, 5:00 PM to 7:00 PM, and 8:00 PM to 10:00 PM. After receiving the notification, participants had a one-hour window to complete the survey and received up to three reminders before the survey expired. Before starting the EMA protocol, participants underwent training on using the mobile application and completed a practice survey in person under the supervision of the research team. They also received follow-up calls on the first two days and periodically thereafter to address any potential questions and concerns.

### C. EMA Design

The EMA survey used in this study was adapted from a previously validated survey designed to measure daily functioning and symptoms in individuals with HIV [24], [25]. The adapted version has demonstrated ecological validity and sensitivity in assessing mental and physical symptoms post-stroke [17], [23].

The EMA survey consisted of 30 questions designed to capture PSI, secondary conditions, and daily activity patterns of study participants (see Supplementary Material for the complete list of questions). More specifically, PSI was a subjective measure reflecting the extent to which participants felt isolated from others. Secondary conditions captured somatic and mood-related symptoms, including pain, fatigue, anxiety, and depression. Responses to these questions were rated on a 5-point Likert scale ranging from 1 (not at all) to 5 (very much). Daily activity patterns captured real-time information about 1) participants’ locations and transportation methods, 2) their social companions, and 3) activities they were engaged in, with responses selected from pre-defined options. In addition, participants were asked to rate their social interactions on a 7-point scale from 1 (not confident/not satisfied/not successful) to 7 (very confident/satisfied/successful) and activities on a 7-point scale from 1 (no help/not well/not satisfied/not engaged) to 7 (a lot of help/very well/very satisfied/very engaged).

### D. Overview of PSI Prediction

Fig. 1 illustrates the data analytic pipeline designed for PSI prediction. In the pre-training phase, language embeddings were first extracted from individual survey questions using a pre-trained language model. The question-level embeddings were concatenated to form the input to an autoencoder. The autoencoder was trained to reconstruct the original input, thereby learning a compact latent representation of the entire survey. In the downstream phase, the pre-trained encoder was leveraged to generate latent representations, which were used as inputs to a supervised classification model for predicting PSI.

**Fig. 1.**
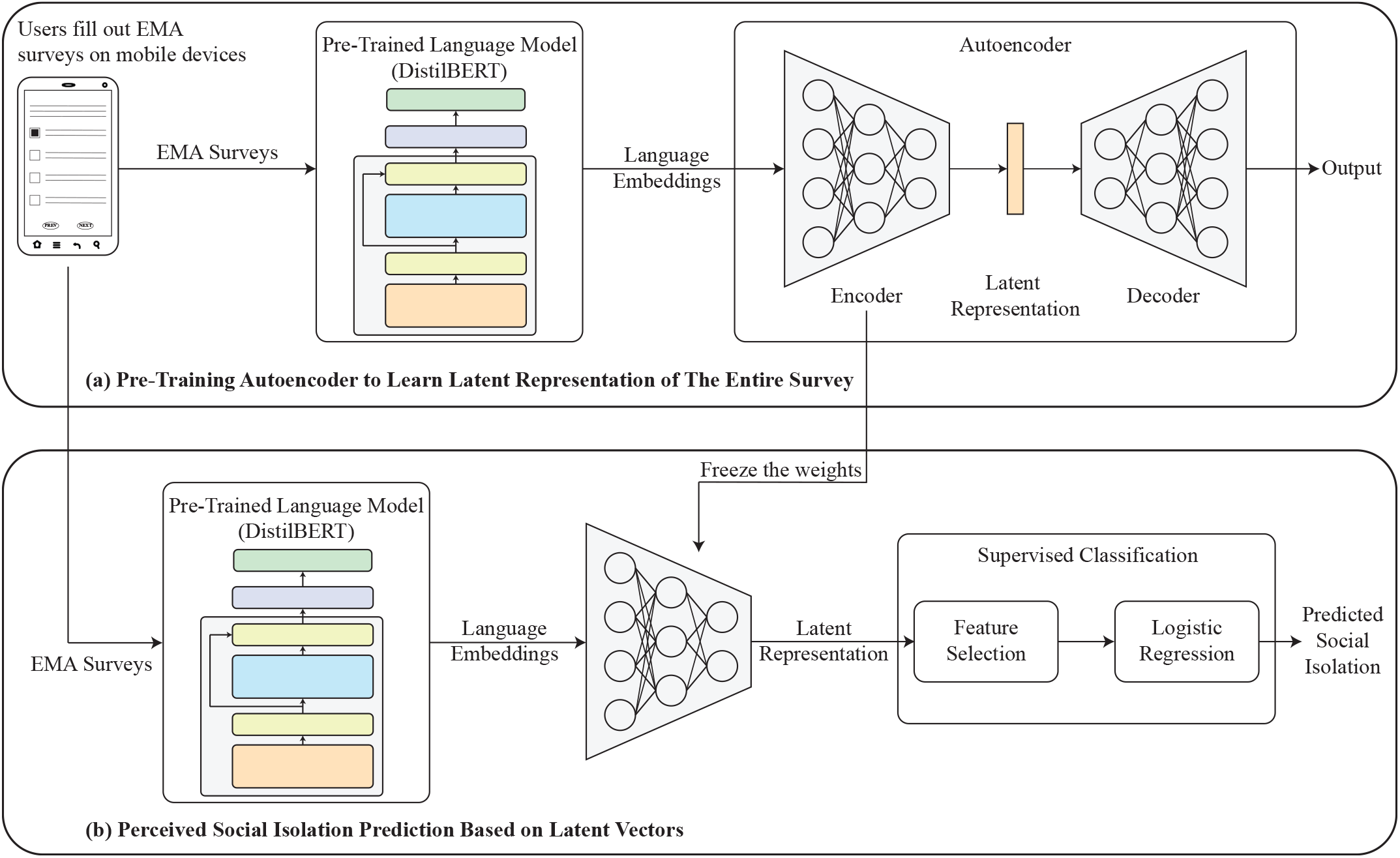
The plots illustrate the data analytic pipeline designed for PSI prediction. (a) Language embeddings were extracted from individual survey responses and concatenated to form the input to an autoencoder. The autoencoder was trained to learn compact latent representations by reconstructing the original input. (b) The pre-trained language model and encoder were then used to generate latent representations of new survey data, which served as inputs to a supervised classification model for predicting social isolation status.

### E. Embedding Extraction and Autoencoder Pre-Training

We utilized pre-trained DistilBERT [26], a lightweight transformer-based model, to extract language embeddings from the EMA data. DistilBERT retains much of the representational capacity of the original BERT model whileenabling faster inference and requiring fewer computational resources. To preprocess the input data, we combined each question prompt with its corresponding patients response into a single textual input. Then, we grouped conceptually related questions, such as those associated with social interactions and activity performance, to provide a more coherent contextual representation, as shown in the example below. In total, 18 textual inputs were generated from the 30 EMA survey questions, and each input was encoded into a 768-dimensional embedding vector.

> *I am at work right now. I drove myself*.
>
> *None of these things have gotten in the way*.
>
> *I have 4 or more interactions with someone else*
>
> *since the last alarm*.
>
> *I am with co-worker(s), friend(s), unknown people*.
>
> *The confidence level in my interaction with the*
>
> *person(s) I am with is 5. The satisfaction level in*
>
> *my interaction with the person(s) I am with is 6. The*
>
> *success level in my interaction with the person(s) I*
>
> *am with is 6*.
>
> *I am doing working (paid). The help I am getting*
>
> *from someone else while doing this activity is 6*.
>
> *The activity performance is 7. The satisfaction in*
>
> *the activity is 7. The engagement in the activity is*
>
> *7*.
>
> *My pain level is mild*.
>
> *I do not feel nervous or stressed at all*.
>
> *I feel a little bit tired*.
>
> *I do not feel depressed at all*.
>
> *I do not feel worthless at all*.
>
> *I have very much trouble concentrating*.
>
> *I do not have difficulty learning new tasks or instructions at all*.
>
> *I do not feel uneasy at all*.
>
> *It is a little bit easy for me to keep track of my*
>
> *thoughts and feelings*.
>
> *I have somewhat little interest in doing things*.
>
> *I do not have a poor appetite at all*.
>
> *My moving/speaking is so slow or restless that other*
>
> *people have noticed not at all*.
>
> *I feel very much cheerful*.

To merge the embeddings from different questions and represent each EMA survey as a single fixed-length vector, we designed an autoencoder to encode the sequence of embeddings. The autoencoder aimed to reconstruct the original input at its output [27], thereby learning a compact latent representation that reflects the underlying semantic structure and interrelationships among the individual survey questions. To prepare the input to the autoencoder, the language embeddings were concatenated and represented as a vector of size 18 *×* 768, corresponding to 18 textual inputs encoded into 768-dimensional DistilBERT embeddings. The encoderconsisted of three fully connected layers with sizes of 9 ×768, 4 × 768, and 2 × 768, respectively. The output of the encoder was a latent representation with 768 dimensions. The decoder mirrored the encoder architecture, consisting of three fully connected layers that reconstructed the original input from the latent representation.

During the training of the autoencoder, data from ten subjects were randomly assigned to the test set, another ten subjects were selected for the validation set, and the remaining 198 subjects were used for training. We employed cross-validation to ensure that each subject appeared in the test set exactly once. The autoencoder was trained to minimize the reconstruction error between the DistilBERT embeddings and the autoencoder output, using mean squared error (MSE) as the loss function. The optimal learning rate for the Adam optimizer was chosen using a logarithmically-spaced grid search [10^*−*4^, 10^*−*2^] based on the performance of the validation set. The autoencoder was trained for up to 50 epochs with early stopping to prevent overfitting. After training, only the encoder was used to generate latent representations for the subsequent PSI prediction.

### F. Social Isolation Prediction

To train a classification model for predicting PSI, we leveraged multiple previous EMA surveys. This approach was motivated by the time-varying nature of PSI, which requires observing participants over time to improve prediction accuracy. Specifically, to predict the PSI level reported at time point *t*_*i*_, surveys collected at time points (*t*_*i−*1_, *t*_*i−*2_, …, *t*_*i−N*_) were used as input, where 1 ≤ *N ≤* 5. A maximum of five previous surveys was used to ensure that predictions could be generated within a day, minimizing delays in potential interventions. Participants rated their PSI level on a 5-point Likert scale, where 1 indicated “not at all” and 5 indicated “very much”. To simplify the prediction task, responses were binarized: a rating of 1 was labeled as negative, indicating “not isolated”, while ratings from 2 to 5 were labeled as positive, indicating “isolated”.

We employed a nested cross-validation strategy to predict whether a participant would experience PSI. In the outer loop of cross-validation, the ten participants designated as the test set during autoencoder pretraining (as described in Section II-E) were kept in the test set, while the remaining participants were put in the training set. This setup ensured that the test participants were not involved in any part of the model training process, thereby preserving generalizability. The input data were the latent representations generated by the autoencoder. These representations were normalized by subtracting the mean and dividing by the standard deviation. To identify informative dimensions within the latent space, we computed the Area Under the Precision-Recall Curve (AUPRC) for each dimension on the training set and selected those with AUPRC greater than a threshold *δ*. For classification, we employed Logistic Regression with L1 penalty to reduce overfitting. Hyperparameters for feature selection and classification were tuned in an inner three-fold cross-validation. Specifically, the AUPRC threshold *δ* was searched uniformly within the range [0.70, 0.85] [28], [29], and the regularization parameter *C* for Logistic Regression was searched logarithmically within the range [10^*−*3^, 10^3^]. Model performance was evaluated using *F*_1_ score. To address class imbalance, *F*_1_ score was computed with sample weights assigned inversely proportional to class distribution. After identifying the optimal hyperparameters, the final model was evaluated on the held-out test participants in the outer loop.

### G. Comparative Analysis

To establish a baseline for comparison, we designed hand-crafted features based on the responses to the EMA surveys. Specifically, ordinal responses were encoded using their original numerical values, with larger values indicating a greater intensity or frequency of the measuring construct. For nominal questions, responses were one-hot encoded into binary features, where a value of one indicated the selected response and zero indicated otherwise. The question “What activity were you doing?” included a large number of possible responses. We reduced complexity by grouping the predefined activities into seven broader categories: activities of daily living (ADL), instrumental activities of daily living (IADL), passive leisure activity, cognitively stimulating activity, physical activity, social activity, and vocational activity [23]. Each category was then represented as a separate binary feature, indicating whether the participant engaged in an activity from that category.

Because multiple EMA surveys could be used for PSI prediction, we aggregated features across up to five surveys to capture temporal patterns, consistent with the proposed method. We computed the mean, standard deviation, maximum, and minimum values for each feature across the available surveys, as suggested in prior work [18]. Additionally, previous studies have shown that temporal dynamics vary across individuals and can provide insight into intra-individual changes. Therefore, we also included variability-related measures, such as the root mean square of successive differences (RMSSD) [30], coefficient of variation [31], and mean successive variability [31]. In total, 567 features were extracted. The model trained on hand-crafted features followed the same procedure described in Section II-F.

## III. RESULTS

### A. Results of PSI Prediction

A total of 11,802 EMA surveys were collected, with 1,707 (14.46%) indicating the presence of PSI. At the participant level, 124 stroke survivors (56.88%) reported experiencing PSI at least once. Although PSI occurred intermittently across individual surveys, its occurrence among more than half of the participants highlights its prevalence and the importance of detecting and addressing social isolation in stroke survivors. Table I summarizes the performance of our language embedding-based PSI prediction using varying numbers of EMA surveys as input. To address class imbalance, precision score, *F*_1_ score, and AUPRC were computed with sample weights inversely proportional to the class distribution. Model performance generally improved with the inclusion of more surveys, indicating that aggregating information across multiple time points contributes to higher prediction accuracy. Performance stabilized around four surveys, with an *F*_1_ score of 0.84 and an AUPRC of 0.92, as summarized in Fig. 2.

**TABLE 1:** Results OF Social Isolation Prediction Based ON Language Embeddings

| # of Surveys | Recall Score | Precision Score | $F_1$ Score | AUPRC |
| --- | --- | --- | --- | --- |
| 1 | 0.78 | 0.84 | 0.81 | 0.88 |
| 2 | 0.80 | 0.85 | 0.83 | 0.90 |
| 3 | 0.81 | 0.86 | **0.84** | 0.91 |
| 4 | **0.82** | **0.87** | **0.84** | **0.92** |
| 5 | 0.81 | **0.87** | **0.84** | **0.92** |

**Fig. 2.**
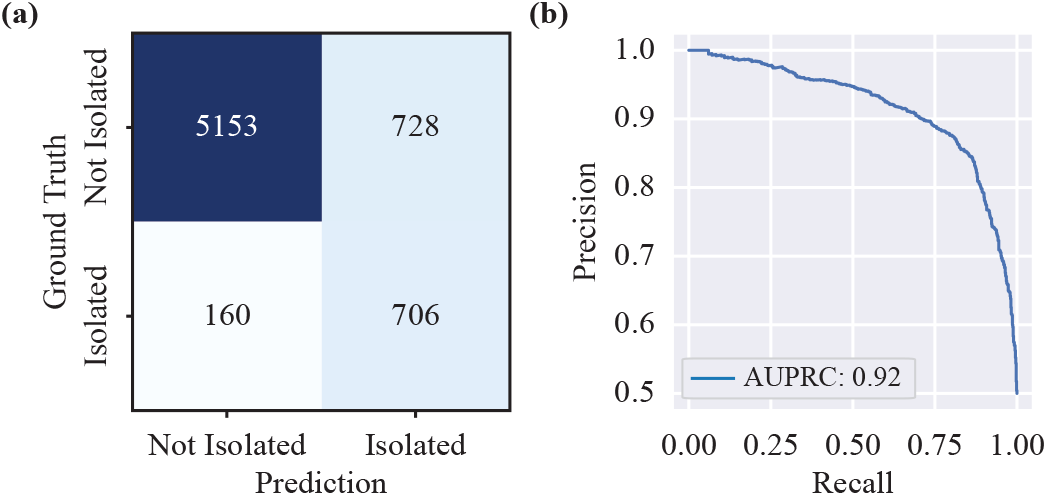
(a) The confusion matrix and (b) Precision-Recall curve of the classification model trained with four EMA surveys as input.

### B. Results of Comparative Analysis

Fig. 3 compares the prediction performances of our proposed language embedding-based model with a model trained on hand-crafted features. Across different numbers of surveys used for training, our model consistently outperformed the hand-crafted feature model. Specifically, with four surveys, our approach achieved a 7% improvement in recall score, 4% improvement in *F*_1_ score, and 4% improvement in AUPRC. These results highlight the effectiveness of representation learning in capturing complex and latent structures within EMA responses that manually crafted features may overlook.

**Fig. 3.**
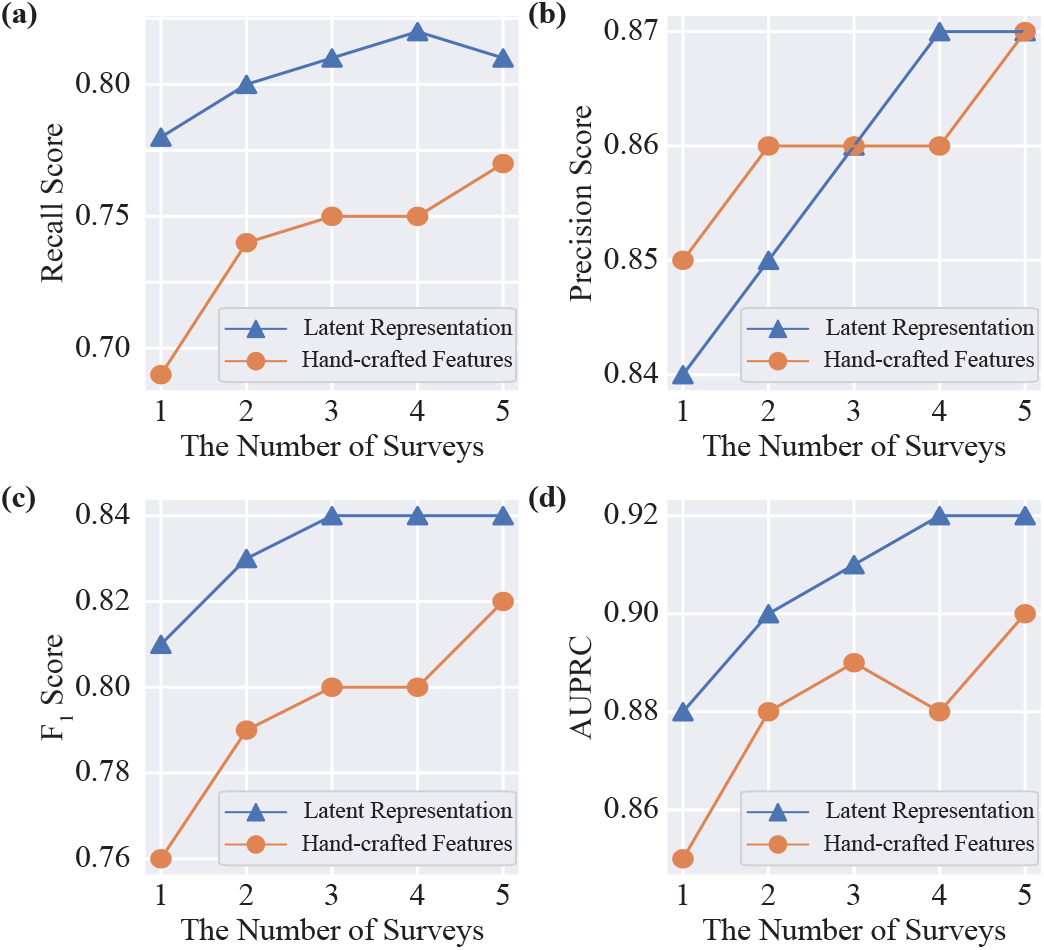
The figures compare the models trained on hand-crafted features and latent representations learned from the autoencoder in terms of (a) recall score, (b) precision score, (c) ***F***_**1**_ score, and (d) AUPRC.

### C. Identify Important Questions in EMA Surveys

We employed an empirical forward selection approach to identify the most informative EMA questions contributing to PSI prediction. Specifically, for each question, we kept only its language embedding and set the embeddings from all other questions to zero. Because the encoder only consisted of fully connected layers, zero-valued embeddings for unselected questions did not contribute to the computation of the latent representations [32]. The modified embeddings were concatenated and passed through the pre-trained encoder to generate latent representation, which was used as input to the downstream classification pipeline. Questions were ranked based on the classification performance in the validation set within the inner cross-validation loop. The question that yielded the best performance was selected first. Subsequently, the question that—when combined with the previously selected question(s)—yielded the most significant improvement was added. This iterative process continued until all questions had been evaluated.

Fig. 4 presents the classification performance achieved by incrementally adding questions through the forward selection approach across different numbers of EMA surveys. Overall, performance improved with both the inclusion of more surveys and the addition of more questions. Notably, substantial improvements in AUPRC were observed with the first few selected questions. For example, using five surveys and only three questions yielded an AUPRC of 0.91, comparable to the performance obtained using all questions (0.92). These results suggest that a small subset of carefully selected questions can provide comparable predictive performance, especially when multiple surveys are available.

**Fig. 4.**
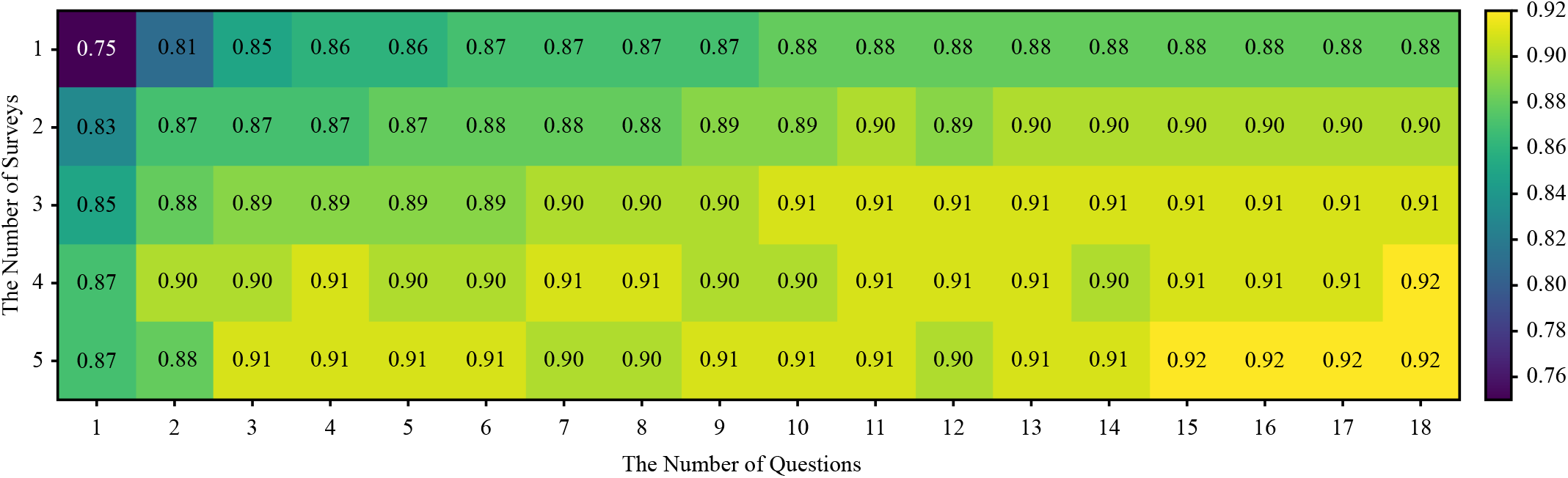
The heatmap illustrates how the classification performance changes with varying numbers of EMA surveys and questions included

When using five surveys and only three questions, the most frequently selected questions in sequential order were 1) levels of anxiety, 2) feelings of worthlessness, and 3) the number of social interactions since the last EMA prompt. Due to the limited interpretability of raw language embeddings, we examined the average self-reported values for these questions across the five surveys, as illustrated in Fig. 5. The Mann–Whitney U test indicated statistically significant differences in the distributions of all three questions between the socially isolated and non-isolated groups (*p <* 0.05). Specifically, participants in the socially isolated group reported higher levels of anxiety, worse feelings of worthlessness, and fewer social interactions compared to their non-isolated counterparts. Although the average number of social interactions showed a statistically significant difference between groups, handcrafted features based on this question did not contribute to the prediction of PSI. However, incorporating the language embedding from this question improved model performance, suggesting that the proposed language embedding-based approach can capture subtle contextual inter-question dependencies that are not reflected in the handcrafted feature values.

**Fig. 5.**
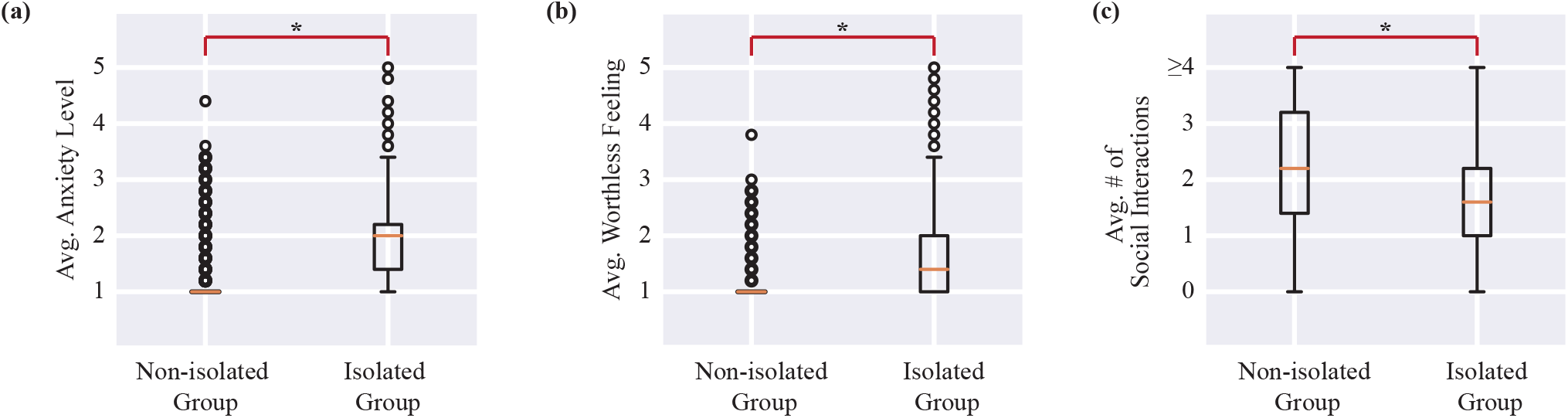
Boxplots of the most important questions for social isolation prediction. The asterisks indicate a significant statistical difference (***p <* 0.05**) in the distribution between the socially isolated and non-isolated groups. Abbreviation: avg.—average

## IV. DISCUSSION

In this paper, we propose a data analytic pipeline for predicting PSI based on language embeddings derived from EMA surveys. An autoencoder was trained to integrate embeddings from participants’ survey responses and produce a compact latent representation of the entire survey. Our results demonstrate that these latent representations captured rich and meaningful information, leading to an improved prediction performance compared to baseline models based on handcrafted features. These findings highlight the effectiveness of representation learning in capturing nuanced patterns from participants’ self-reported data.

Previous studies on social isolation have primarily focused on identifying factors associated with loneliness and social well-being [33]–[38]. While these studies offer promising insights, their primary objective was not to develop machine learning models to predict social isolation in dynamic, real-world settings. Without validation in predictive contexts, the practical utility of their findings remains limited. A few studies have explored predictive modeling for the automatic detection of social isolation. For instance, Martinez *et al*. used features related to communication, mobility, and demographics to predict social isolation among older adults, reporting an accuracy of 100% [39]. However, this result was based on a small sample of only seven participants, which may offer limited generalizability. Doryab *et al*. examined the use of passive sensing data to infer loneliness levels among college students, achieving an *F*_1_ score of 0.80 [40], notably lower than our approach. Moreover, both studies were conducted in healthy populations whose physical, cognitive, and psychosocial characteristics differ significantly from those of stroke survivors. As a result, the findings may not generalize well to post-stroke populations.

We identified a small subset of EMA survey questions that significantly contributed to predicting social isolation, which is consistent with previous findings. The first important question was related to anxiety. Santini *et al*. reported a bidirectional relationship between social isolation and anxiety [41]. More specifically, individuals who lack social support are at increased risk of depression and anxiety, while elevated anxiety can lead to social withdrawal, further deepening feelings of loneliness and PSI. This dynamic is particularly relevant for stroke survivors, who often face a combination of physical limitations and insufficient social support. Notably, approximately 45% individuals with stroke report feeling abandoned after hospital discharge [17]. In addition, feelings of worthlessness have also been linked to PSI. A qualitative study by Freeman *et al*. examined individuals with multiple sclerosis and found that limited physical ability contributed to feelings of guilt and worthlessness, which in turn reinforced social isolation [42].

These psychological and social dynamics may extend to stroke survivors, who often face similar physical limitations and emotional challenges. A recent post-stroke EMA study, which utilized network analysis to examine the original EMA survey responses, found that feelings of worthlessness, both temporally and contemporaneously, predict PSI [17]. This study, along with our project, indicates that stroke survivors are more likely to report feelings of isolation in follow-up surveys when they have previously reported feeling worthless. Experiencing worthlessness can hinder individuals from forming and maintaining meaningful social relationships, which can lead to a decline in social interactions and an increased feeling of isolation [43]. Finally, the number of recent social interactions was also identified as an important factor. This factor is straightforward, as limited social participation is a common consequence of a stroke. Research shows that stroke survivors often develop very small social networks, with their most significant relationships primarily involving immediate family members [44]. Fewer social interactions suggest higher levels of social disconnectedness, which research has identified as a distinct form of isolation that negatively impacts psychosocial outcomes [45].

We envision a scenario where EMA surveys are deployed on mobile devices with only a small subset of informative questions. We aim to optimize the trade-off between predictive accuracy and user engagement by limiting the survey to as few as three items. Once responses are submitted, the predicted PSI status will be identified and displayed immediately. If the system detects a risk of social isolation, it can trigger timely interventions for participants to reduce this isolation.

This work has several limitations. First, participants answer the EMA survey based on predefined response options, which may not fully capture personal feelings and experiences. Future work could explore the opportunity for patients to type about their current emotional state in short free-text or voice-based natural language formats. This approach could provide richer contextual data, capturing nuances that might be missing in structured surveys. Second, although our model achieved strong predictive performance, it relies on self-reported data, which may be subject to individual biases or inconsistencies. Future work could incorporate more objective, multi-modal sensor data, such as inertial, heart rate, skin temperature, and bio-impedance, to complement subjective responses and provide a more comprehensive understanding of the individual’s condition.

## V. CONCLUSION

This paper demonstrates that language embeddings extracted from structured EMA surveys contain valuable information for predicting PSI. By leveraging only a small subset of questions, a trade-off between validity and usability can be achieved. These findings highlight the potential of scalable, low-burden digital tools for real-world deployment, supporting continuous mental health monitoring and enabling more personalized rehabilitation strategies for stroke survivors.

## Data Availability

All data produced in the present study are not available

## ACKNOWLEDGMENT

We would like to thank Dr. Jie Xiong and Dr. VP Nguyen for their valuable feedback on this work.

