## Supplemental materials for "Leveraging Language Embeddings from EMA Surveys to Predict Perceived Social Isolation among Stroke Survivors"

### Supplementary Materials

Table S1: Survey Questions from The Ecological Momentary Assessment (EMA) Used in This Study

| Questions | Description | Response Options |
|---|---|---|
| where_are_you | Where are you right now? | 1.At my home<br>2.At the home of family members(s)<br>3.At the home of friend(s)<br>4.At work<br>5.At outpatient medical visit<br>6.In hospital<br>7.At restaurant<br>8.At community center<br>9.In public business/store (e.g., post office, grocery store)<br>10.In a vehicle<br>11.Outside, walking<br>12.In class/school setting<br>13.Inside, other<br>14.Outside, other |
| how_get_where_are Answer the question only if where_are_you!=1 | How did you get where you are? | 1.Walked<br>2.Got a ride from someone<br>3.Took bus or train<br>4.Drove myself<br>5.Other |
| environmental_barriers | Since the last alarm, have any these things limited your gotten in the way? (Check all that apply) | 1.None<br>2.Built Environment barriers (e.g., no curb cuts, no accessible ramp)<br>3.Weather (e.g., temperature, humidity, rain, wind, air quality)<br>4.Lights (e.g., overly bright lights, flashing lights, low lighting, or darkness)<br>5.Sounds (e.g., loud noises)<br>6.Difficulty reading or understanding written materials<br>7.Difficulty understanding what someone is saying<br>8.People's attitudes<br>9.Traffic, parking, or transportation problems<br>10.Other |

| | | |
|---|---|---|
| times_socialized | Since the last alarm, how many times have you interacted with someone else? | 1.0 (no interactions)<br>2.1 interaction<br>3.2 interactions<br>4.3 interactions<br>5.4 or more interactions |
| who_is_with_you | Who is with you right now? (Check all that apply) | 1.Alone<br>2.Co-worker(s)<br>3.Family member(s)<br>4.Friend(s)<br>5.Healthcare provider(s)<br>6.Pet(s)<br>7.Spouse or partner<br>8.Other known people<br>9.Unknown people<br>10.People you are socializing with electronically |
| confidence_social<br>Answer the question only if who_is_with_you!=1 and who_is_with_you!=6 | I am confident in my interaction with the person(s) I am with... | slider<br>1, Not confident, 7, Very confident |
| satisfaction_social<br>Answer the question only if who_is_with_you!=1 and who_is_with_you!=6 | I am satisfied with my social interaction... | slider<br>1, Not satisfied, 7, Very satisfied |
| success_social<br>Answer the question only if who_is_with_you!=1 and who_is_with_you!=6 | I am successful with my social interaction... | slider<br>1, Not successful, 7, Very successful |
| why_not_engaged_social<br>Answer the question only if who_is_with_you=1 or who_is_with_you=6 | I am not doing anything social because... (Check all that apply) | 1.I'm busy<br>2.I don't have the opportunity<br>3.I'm happy being alone (no motivation for social interactions)<br>4.I don't have the means (e.g., money, transportation)<br>5.I am in pain<br>6.I want to but I'm tired<br>7.Other |

| | | |
|---|---|---|
| what_doing_home<br>Answer the question only if where_are_you=1 | What are you doing? | 1.Arts and crafts<br>2.Budgeting or paying bills<br>3.Care of others<br>4. Changing clothes<br>5.Doing household chores (e.g., laundry, cleaning)<br>6.Eating or drinking at home<br>7.Exercising<br>8. Gardening<br>9.Household projects/car maintenance<br>10.Internet/computer/tablet use<br>11.Listening to music/radio<br>12.Looking for a job<br>13.Mediating<br>14.Playing a musical instrument<br>15.Playing with children<br>16.Playing with pets<br>17.Preparing food/cooking<br>18.Private religious activities<br>19.Reading, writing, or journaling<br>20.Resting<br>21. Schoolwork<br>22. Shopping online<br>23.Showering or grooming<br>24.Social media (e.g., Facebook, Twitter)<br>25.Social interactions with someone<br>26.Smoking<br>27.Talking on the phone/texting<br>28.Watching TV<br>29.Working (paid)<br>30.Working (unpaid) or volunteering<br>31.Other physical activity<br>32.Other social activity<br>33.Other mental stimulating activity<br>34.Nothing |

| | | |
|---|---|---|
| doing_not_home<br>Answer the question only if where_are_you!=1 | What are you doing? | 1.Caring for others<br>2.Dong laundry away from home<br>3. Eating or drinking out<br>4. Entertainment (cinema, sports, etc.)<br>5. Exercising<br>6.Getting gas<br>7.Household projects/car maintenance<br>8. Internet/computer.tablet use<br>9.Listening to music/radio<br>10.Looking for a job<br>11.Meditating<br>12.Meeting (church, parent group, AA, etc.)<br>13.Participating in a social event (party, wedding, etc.)<br>14.Playing with children<br>15.Playing with pets<br>16.Reading, writing, journaling<br>17.Private religious activities<br>18.Resting<br>19.Riding in a bus, trolley, car, or van<br>20.Schoolwork<br>21. Shopping outside of the home<br>22. Smoking<br>23.Social interactions with someone<br>24.Social media (Facebook, Twitter)<br>25.Talking on the phone/texting<br>26.Traveling<br>27.Visiting the beach, park, etc.<br>28.Visiting a museum, concert hall, etc.<br>29. Visiting the barber/hairdresser/nail salon<br>30.Visiting family or friends<br>31. Visiting healthcare providers (doctors, nurses, therapists)<br>32.Watching TV<br>33.Working (paid)<br>34.Working (unpaid) or volunteering<br>35.Other physical activity<br>36.Other social activity<br>37.Other mental stimulating activity<br>38.Nothing |

| | | |
|---|---|---|
| help<br>Answer the question only if what_doing_home!=20 and what_doing_home!=34 and doing_not_home!=18 and doing_not_home!=38 | How much help are you getting from someone else while doing this activity? | slider<br>1, No help, 7, A lot of help |
| perform_activity<br>Answer the question only if what_doing_home!=20 and what_doing_home!=34 and doing_not_home!=18 and doing_not_home!=38 | I am performing this activity well... | slider<br>1, Not well, 7, Very well |
| satisfied_activity<br>Answer the question only if what_doing_home!=20 and what_doing_home!=34 and doing_not_home!=18 and doing_not_home!=38 | I am satisfied with doing this activity... | slider<br>1, Not satisfied, 7, Very satisfied |
| engaged_activity<br>Answer the question only if what_doing_home!=20 and what_doing_home!=34 and doing_not_home!=18 and doing_not_home!=38 | I am engaged in doing this activity... | slider<br>1, Not engaged, 7, Very engaged |

| | | |
|---|---|---|
| why_not_doing_anything Answer the question only if what_doing_home=20 or what_doing_home=34 or doing_not_home=18 or doing_not_home=38 | Why aren't you doing anything? (Check all that apply) | 1.I'm satisfied with doing nothing at this time<br>2.I am in pain<br>3.I am tired<br>4.I'm feeling down or sad<br>5.I don't know what to do<br>6.I don't have someone to do something with<br>7.I don't have the means to do something (e.g., money or transportation)<br>8.I can't overcome physical barriers (e.g., weather, no accessible ramp)<br>9.I can't overcome negative attitudes of people toward my disability<br>10.Other |
| pain | Right now, my pain level is... | 1.No pain<br>2.Mild<br>3.Moderate<br>4.Severe<br>5.Very Severe |
| isolated | Right now, I feel isolated from others... | 1.No at all<br>2.A little bit<br>3.Somewhat<br>4.Quite a bit<br>5.Very much |
| stress | Right now, I feel nervous and stressed... | 1.No at all<br>2.A little bit<br>3.Somewhat<br>4.Quite a bit<br>5.Very much |
| tired | Right now, I feel tired... | 1.No at all<br>2.A little bit<br>3.Somewhat<br>4.Quite a bit<br>5.Very much |
| depressed | Right now, I feel depressed... | 1.No at all<br>2.A little bit<br>3.Somewhat<br>4.Quite a bit<br>5.Very much |

| | | |
|---|---|---|
| worthless | Right now, I feel worthless... | 1.No at all<br>2.A little bit<br>3.Somewhat<br>4.Quite a bit<br>5.Very much |
| concentrating | Right now, I have trouble concentrating... | 1.No at all<br>2.A little bit<br>3.Somewhat<br>4.Quite a bit<br>5.Very much |
| learning_new | Right now, I have difficulty learning new tasks or instructions... | 1.No at all<br>2.A little bit<br>3.Somewhat<br>4.Quite a bit<br>5.Very much |
| anxiety | Right now, I feel uneasy... | 1.No at all<br>2.A little bit<br>3.Somewhat<br>4.Quite a bit<br>5.Very much |
| mindfullness | Right now, it is easy for me to keep track of my thoughts and feelings... | 1.No at all<br>2.A little bit<br>3.Somewhat<br>4.Quite a bit<br>5.Very much |
| little_interest | Right now, I have little interest in doing things... | 1.No at all<br>2.A little bit<br>3.Somewhat<br>4.Quite a bit<br>5.Very much |
| appetite | Right now, I have a poor appetite or I am overeating... | 1.No at all<br>2.A little bit<br>3.Somewhat<br>4.Quite a bit<br>5.Very much |
| slow_restless | Right now, my moving/speaking is so slow or restless that other people have noticed... | 1.No at all<br>2.A little bit<br>3.Somewhat<br>4.Quite a bit<br>5.Very much |

| | | |
|---|---|---|
| cheerful | Right now, I feel cheerful... | 1.No at all<br>2.A little bit<br>3.Somewhat<br>4.Quite a bit<br>5.Very much |
